# Early Childhood Reading for Pleasure: Evidence from the ABCD Study for Benefits to Cognitive Performance and Mental Health and Associated Changes in Brain Structure

**DOI:** 10.1101/2022.02.27.22271550

**Authors:** Yunjun Sun, Barbara J. Sahakian, Christelle Langley, Anyi Yang, Yuchao Jiang, Xingming Zhao, Chunhe Li, Wei Cheng, Jianfeng Feng

## Abstract

**BACKGROUND:** Early childhood has increasingly been recognized as an important neurodevelopmental period. However, the links between specific forms of early educational activities and cognition and mental health remain unclear.

**METHODS:** A large-scale analysis on the attainments of childhood Reading for Pleasure (early RfP) and its relationship with young adolescent measures of cognition, mental health and brain structure was conducted, using linear mixed-effects model, structural equation and mediation analyses. Participants were from the Adolescent Brain and Cognitive Development (ABCD) study, a USA national cohort (n=11,878). A two-sample Mendelian randomization (MR) analysis using genetic instruments from two independent genome-wide association studies (GWAS) based on ABCD and GWAS meta-analyses including UK-biobank datasets were conducted to assess potential causal inference.

**RESULTS:** Early RfP was positively associated with cognitive assessments, which in contrast, was negatively associated with dimensional psychiatric problems in young adolescents. MR analysis revealed a beneficial relationship between early RfP and later-life cognitive performance, and a trend towards a protective relationship between early RfP and later-life attention disorder. Brain regions in which larger cortical areas were positively associated with early RfP included the superior temporal, prefrontal, left angular, parahippocampal, right anterior cingulate (ACC), occipital, supramarginal and orbital regions and subcortical ventral diencephalon (DC), thalamus, and brainstem volumes. Mediation analysis indicated that the brain regions significantly mediated the effects of early RfP with better cognitive performance and lower psychiatric problems.

**INTERPRETATION:** Our results highlight the importance of encouraging RfP in young children during the critical early childhood stage, as it was associated with beneficial outcomes for cognition, mental health, and brain structure later in life.

## Introduction

Early childhood is a critical period for brain development and plasticity in learning to foster good cognition and better mental health and well-being throughout life(1, 2). The benefits of good brain health and the development and establishment of behaviour in childhood that promote this is key to healthy development and mental well-being into adolescence and adulthood, providing resilience in times of stress(3). It is now known that our brains are in development during childhood and adolescence. Neuroimaging studies revealed that growth in the brain cortical grey matter volume (GMV) is greatest during the first 2 years of life, and the surface area expands until approximately 8-12 years of age, after which it declines progressively and there simultaneously tends to be a gradual decrease in the GMV and cortical thickness(4, 5). These later changes may be due in part to subsequent pruning of synaptic connections and functional specialization(5-7). Cognitive progression milestones parallel the sequence of brain maturation: Brain regions associated with primary functions, including the motor and sensory systems, are the earliest to mature; subsequently, regions linked to basic language and spatial attention functions mature, including the temporal and parietal association regions, with the developmental curve starting to peak at 9 months after birth(5, 8, 9). Studies have indicated the importance of the early stages of learning, including learning language(10). Similar to adult language processing, infants achieve complex, hierarchical organization of the brain during language acquisition(11). In addition, they can learn one or multiple languages to which they are exposed, simultaneously acquiring languages through either auditory-vocal or visual-manual modes (12), though a reduction in this initial-language learning flexibility usually occurs with age(10, 13).

Compatible with the rapid rate of brain development and learning in early childhood, evidence was found for the benefits of early educational or intervention initiatives on later well-being. The Child Parent Center Education Program (CPCEP) in Chicago reported persistent benefits for children who began preschool education earlier at 3 to 4 years old compared with those who began kindergarten at an older age. The former preschool participants had higher educational levels and socioeconomic status and lower rates of substance abuse in adulthood(14, 15). A study in England reported that high-quality preschool education had long-term impacts on the literacy and numeracy of children at age 11. The magnitudes of the benefits were equal to or exceeded the effects of some negative factors, including early developmental problems and poorer primary school qualities(15, 16). Furthermore, the particularly importance of early childhood for intervention in one typical neurodevelopmental disorder, autism spectrum disorder (ASD), was also indicated. Behavioural intervention initiated below 30 months of age using the Early-Start-Denver Model (ESDM) sustainably altered the long-term developmental course of autism, with core symptoms improved(17).

These results suggest that the early stage is critical for neurodevelopment, cognition, learning and behaviour. To optimize typical development, it is important to seize the critical period for early educational activities(18) and to make this experience of good quality, thus ensuring the best outcomes in future life.

One important type of early educational and learning activity in young children is Reading for Pleasure (RfP). The concept of early RfP attainments has similarities with educational attainments(19, 20), but it is a more enjoyable activity during the critical early development stage. Fundamentally, reading is a cognitive process of gaining language and information in written form, and as a non-innate ability, it is generally learned through explicit training, which contributes more broadly to the acquisition of skills(21, 22). Moreover, during RfP, children not only practice cognitive phonological/orthographic processing of reading but also enjoy absorbing knowledge and wisdom intently. With assistance from caregivers, young children can learn initial printed languages with illustrations, engage in discussions of the educational images and contents, and enjoy reading. Multiple benefits of early reading activities on future advanced reading and language abilities have been reported(21, 23, 24).

To the best of our knowledge, no previous research has investigated the relationship of early RfP with dimensional cognition and mental health and brain structure. The recent ABCD project, which recruited 11,878 youths, provided an opportunity to conduct such a study(25, 26). Cross-sectional and longitudinal data across multiple domains from young adolescents aged 9 ― 11 were analysed this study. We hypothesized that, in comparison with peers who began RfP quite late or never had RfP, youths who enjoyed RfP during early childhood would exhibit long-term benefits, including improved cognitive performance, a decreased risk of developing psychiatric symptoms, and increased academic achievement during typical-development. We also aimed to examine the underlying brain structural mechanisms and determine whether our hypothesis was mediated by brain structure.

## Materials and Methods

### Participants

Data from the third (for baseline data collected between 2016 ― 2018) and fourth (for follow-up data collected between 2017 ― 2020) updates of the ABCD research project (Release 3.0 and 4.0) were used in this study. The ABCD project (https://abcdstudy.org/) is an ongoing longitudinal study with 21 research sites across the US. Participant recruitment and information are provided in **Method.S1**. The ABCD project conforms to the ethical guidelines of each research site’s Institutional Review Board (IRB). The institutional review committee of the University of California, San Diego (UCSD), is responsible for the ethical oversight of the ABCD research. All participants provided written consent forms (parents/caregivers) and informed consent forms (youths). The demographic characteristics are summarized in **Table.1**.

**Table 1.** Demographic characteristics of all participants included in the ABCD study.

| Variables | N (%) or mean (±SD) | | | |
| --- | --- | --- | --- | --- |
| | Release 3.0<br>Total Participants<br>(n = 11878) | Childhood RfP<br>0-2.5 years (late/never)<br>(n = 4942) | Childhood RfP<br>3-5.5 years (early)<br>(n = 4172) | Childhood RfP<br>≥ 6 years (early)<br>(n = 1129) |
| **Age (months)** | 118.98 (±7.49) | 118.39 (±7.49) | 119.21 (±7.5) | 120.3 (±7.24) |
| **Sex** |  |  |  |  |
| Female | 5,682 (47.84) | 2004 (40.55) | 2251 (53.95) | 614 (54.38) |
| Male | 6,196 (52.16) | 2938 (59.45) | 1921 (46.05) | 515 (45.62) |
| Missing/undefined | 0 (0) |  |  |  |
| **Race Ethnicity** |  |  |  |  |
| Asian | 252 (2.12) | 69 (1.40) | 111 (2.66) | 27 (2.39) |
| Black | 1784 (15.02) | 750 (15.18) | 407 (9.76) | 174 (15.41) |
| Hispanic | 2411 (20.31) | 1150 (23.27) | 664 (15.92) | 148 (13.11) |
| Other | 1247 (10.50) | 504 (10.20) | 432 (10.35) | 150 (13.29) |
| White | 6182 (52.05) | 2469 (49.95) | 2558 (61.31) | 630 (55.80) |
| Missing/undefined | 2 (0.00) |  |  |  |
| **Highest Parental Education** |  |  |  |  |
| ≤ HS Diploma/GED | 1725 (14.52) | 806 (16.30) | 314 (7.53) | 99 (8.77) |
| College and Bachelor | 6095 (51.31) | 2751 (55.67) | 2034 (48.75) | 524 (46.41) |
| Post Graduate Degree | 4043 (34.04) | 1385 (28.03) | 1824 (43.72) | 506 (44.82) |
| Missing/undefined | 15 (0.13) |  |  |  |
| **Family Income** |  |  |  |  |
| < \$50,000 | 3224 (27.14) | 1759 (35.59) | 904 (21.67) | 252 (22.32) |
| ≥ \$50,000 & <100,000 | 3070 (25.85) | 1438 (29.10) | 1182 (28.33) | 314 (27.81) |
| ≥ \$100,000 | 4565 (38.43) | 1745 (35.31) | 2086 (50.00) | 563 (49.87) |
| Missing/undefined | 1,019 (8.58) |  |  |  |

Of the 11,878 participants aged 9-11 with baseline data in ABCD cohort, we included 10,243 participants (86.2%) who had full data recordings for the variable of interest (RfP measurements). While 48.2% of the participants started RfP at quite later age or never started (0-2.5 years of RfP, n=4942), 40.8% of the participants had experienced early childhood RfP (3-5.5 years, n=4172), and the remaining 11% began RfP at very young childhood ( ≥ 6 years, n=1129).
RfP, reading for pleasure; ABCD, adolescent brain cognitive development study.

### RfP measurements

The early RfP scale is based on the ABCD Sports and Activities Involvement Questionnaires(abcd_spacss01) completed by caregivers regarding their children. The following 3 items are related to RfP measurements: **1)** question 1, “For how many years has your child read for pleasure?”, representing attainments of early childhood RfP, which is abbreviated as “early RfP” in the study. A higher score on question 1 indicates that the youth started RfP earlier and spent more formative years enjoying it. 1-year and 2-year follow-up data were available for early RfP, and the question was: “Since we last saw you on [asnt_timestamp_c], has your child read for pleasure?”. **2)** question 2, “Approximately how many hours per week does your child spend reading for pleasure?” (RfP duration/week); and **3)** question 3, “About how many hours per week does your child spend reading for pleasure by bin number?” The latter two questions represent RfP habits in young adolescence. Higher scores for questions 2 and 3 indicate longer RfP durations/week. 2-year follow-up data was available for RfP duration and the question was the same as at baseline. While young children’s RfP can be initiated/inspired and facilitated by caregivers, young adolescent’s RfP is relatively more autonomous.

### Structural neuroimaging data

Quality-controlled high-resolution neuroimaging data processed with FreeSurfer were used. The ABCD imaging parameters and protocols are illustrated in https://abcdstudy.org/images/Protocol_Imaging_Sequences.pdf. Neuroimaging data were obtained using 3T scanners with a 32-channel head coil and high resolution T1-weighted structural magnetic resonance imaging (MRI). Both the methods and evaluations of these MRI images have been harmonized and optimized across all ABCD research sites(25, 27). According to the ABCD standard protocol and pipeline, preprocessed T1 sMRI data were obtained. The preprocessing processes were completed by the ABCD research teams, with details described in their image processing paper(27). Whole-brain and regional morphometric structure assessment values containing cortical volume, thickness, and surface areas were obtained using FreeSurfer processing, including 148 cortical regions from the Destrieux atlas(28), 68 cortical regions from the Desikan/Killiany atlas(29), and 40 subcortical segmentations from the ASEG atlas(30), which were used to conduct additional analysis of the association between brain structures and RfP measurements. **Method.S2** contains additional information on sMRI.

The quality-control (QC) procedure of the processed neuroimages was checked by the ABCD team(27) both automatically and manually (by sampling). According to the FreeSurfer reconstruction QC measures(freesqc01), 529 participants who failed QC were removed from structural analyses.

### Assessments of cognitive performance and mental problems

An extensive battery of clinical interviews, including self- and parent-reports, and cognitive and psychiatric tests were completed by both the children and their parents/caregivers. For cognitive performance, we analysed the standard scale of the well-validated National Institutes of Health (NIH) Toolbox (TB) (abcd_tbss01), which was designed to harmonize data collection across NIH-funded projects and to facilitate cross-study comparisons. It contains 2 standardized indices of crystallized (acquired knowledge) or fluid composites of cognition that are comparable to widely used IQ measures(31). Additionally, it evaluates the cognition of the following 7 major components(32): 1). dimensional change card sort, 2). flanker inhibitory control and attention, 3). list-sorting working memory, 4). oral reading recognition, 5). pattern comparison processing speed, 6). picture vocabulary and 7). picture sequence memory, and total score. Details are listed in **Table S1**.

Psychiatric problems were mainly evaluated based on the Child Behaviour Checklist (CBCL) scale (abcd_cbcls01) completed by parents/caregivers. It contains the following 20 experience-based symptoms/problems subscales modified from the CBCL(33) and applied to ABCD youths: 1). ADHD symptom CBCL-DSM5, 2). aggressive, 3). anxious/depressed, 4). anxious problem CBCL-DSM5, 5). attention problem, 6). conduct problem CBCL-DSM5, 7). depression symptom CBCL-DSM5, 8). rule-breaking, 9). obsessive-compulsive CBCL-OCD, 10). internalizing and 11). externalizing broad band scores, 12). oppositional CBCL-DSM5, 13). Sluggish Cognitive Tempo, 14). somatic complaints, 15). somatic problems CBCL-DSM5, 16). stress, 17). social, and 18). thought problems, 19). withdrawn/depressed, and 20). total problems. These were derived from the Parent General Behaviour Inventory for Children and Adolescents(34) and the Prodromal Questionnaire(35). Details are listed in **Table S1**.

### Mendelian randomization (MR) analysis

To evaluate the potential causal associations between early RfP and later-life cognition and attention problems, a two-sample MR analysis was conducted using the ‘‘TwoSampleMR’’ package(36, 37), with analyses performed in R4.0.3. The MR method requires that all genetic variants be valid instrumental variables based on the MR assumptions (**Figure S1**, modified from Choi et al., 2019(37)). Details of the exposure/outcome data are provided in **Method S3**.

The exposure and outcome data containing the following indices were harmonized using the ‘‘TwoSampleMR’’ package, which included single-nucleotide polymorphisms (SNP)s, *P* values, effect alleles (with frequencies), reference alleles, estimated effect sizes (using β statistics if the outcomes were continuous, e.g., cognitive performance scores, and converting Odds-Ratio (OR) to β statistics by log-transformation if the outcomes were binary, e.g., ADHD case–control), and standard errors (SE)s. For each direction of potential causal association, the MR estimates were combined using inverse variance weighted (IVW) meta-analysis, which was equivalent to a weighted regression of the SNP-outcome coefficients on SNP-exposure coefficients to make an overall estimate of causal effect, while the intercept was constrained to zero(37).

We then compared IVW results with two other established MR methods, including 1) a weighted median analysis that allows for half of the instrument variables to be invalid in the causal estimation(38) and 2) the MR–Egger regression with its intercept representing average pleiotropic bias and slope representing the causal estimate(39), which are recognized as being more robust to horizontal pleiotropy but at the expense of decreased statistical power(40). The Steiger-directionality test was further applied to test the causal direction between the hypothetical SNP exposures and SNP outcomes(41)

For sensitivity analyses, the MR-PRESSO (MR_Pleiotropy_Residual_Sum_and Outlier) test was applied to detect pleiotropic outliers(42). Horizontal pleiotropy and heterogeneity were also tested using the MR–Egger intercept test and modified Q-statistics(36).

### Statistical analysis

#### Association study

Linear mixed-effects model (*LMM*) analyses were applied to identify the relationships between early RfP, brain structure, and a wide range of behavioural assessments, including cognition and psychiatric scores. The *LMM* model was recommended by the ABCD research consortium and has been used in several related studies(43-45) to consider the correlated observations within family structures due to siblings and twins and at different sites. In this study, *LMM* was performed using the *fitlme* function in MATLAB (R2019b, MathWorks), which was specialized to model families nested within sites. A participant’s data were excluded if the early RfP information was missing. Generally, in the *LMM* model, the dependent variables were scores related to cognitive performance, psychiatric problems or brain structural measurements; the fixed effects were the early RfP/RfP durations; and the nuisance covariates and random effects were the family structures nested within different sites. Other covariates that were controlled for *LMM* included age, sex, BMI, puberty, race/ethnicity of youths, and family SES, including parental education and family income. For brain structure related analysis, the different types of MRI scanners were also added as covariates. Parental education was defined by the highest education level achieved by both parents. Early RfP, age, BMI, family income and parental education were continuous factors. Sex, race/ethnicity and MRI scanners were categorical factors. If the values of family SES or participants’ demographic information (including sex, age, and race/ethnicity) were not provided (recorded as “Don’t know” or “Refuse to answer”), the missing values were replaced with nan. Details of the *LMM* model are shown in **Method S4**.

#### Longitudinal analysis and Mediation study

A classic two-wave cross-lagged panel model (CLPM) was conducted to perform the longitudinal analysis of the RfP measurements with cognitive assessments, and ADHD symptom scores, which was implemented using Mplus 7.4. Subsequently, we conducted the standard mediation analysis using the Mediation toolbox developed by Tor Wager’s team (https://github.com/canlab/MediationToolbox). Details are shown in **Method S5**.

## Results

### The associations of early RfP with cognitive and psychiatric assessments of young adolescents in the ABCD database

As shown in **Figure 1** and **Table S2**, early RfP had significant positive associations with the standard cognitive assessment scales (Bonferroni-corrected *P* < 0.05), including: **1)** the NIH-TB cognition summary for ABCD youth (abcd_tbss01, *r* values ranging from 0.041 to 0.331) and **2)** the verbal-learning and immediate-memory scale (absd_ps01, *r*: 0.035 ― 0.154). Additionally, early RfP was associated with youths’ speech development (dhx01.devhx_21_p, *r =* 0.1402, *P* < 10^−20^) and school achievement (course grades: *r* = 0.201, *P* < 10^−20^; school performance: *r* = 0.267, *P* < 10^−20^) **(Table S3)**, but was less significantly associated with how early they said their first words (dhx01.devhx_19d_p, *r* = 0.044, *P* = 3.68×10^−4^).

**Figure. 1.**
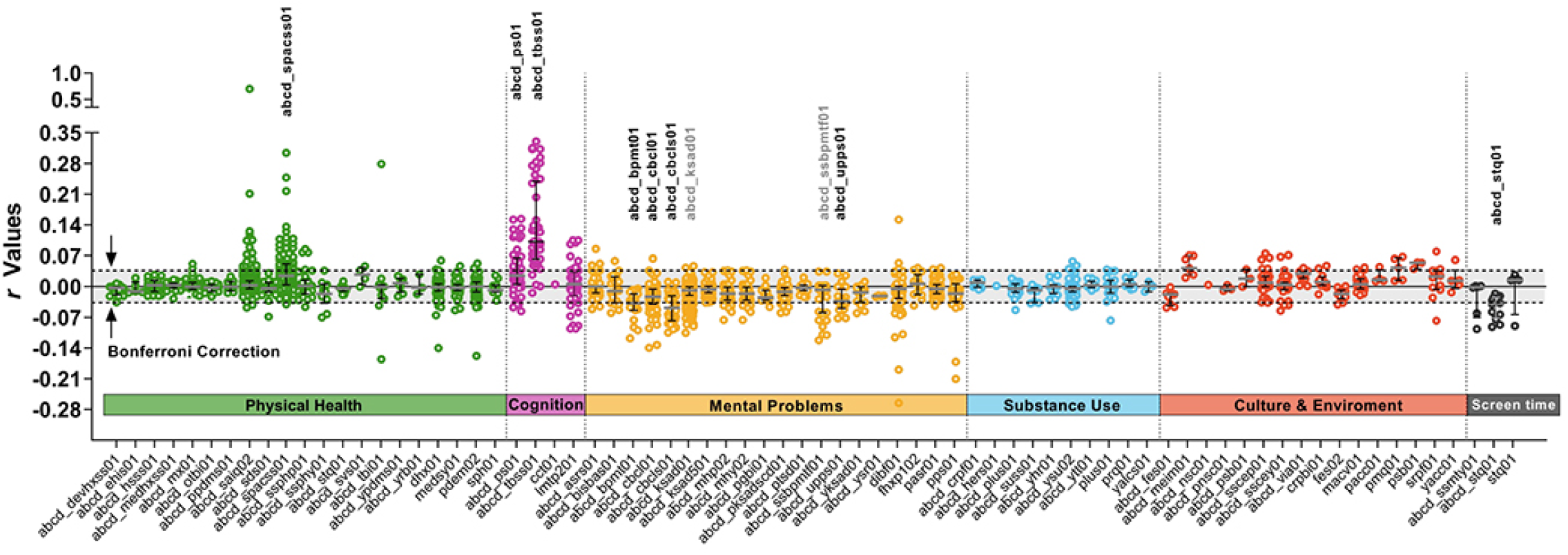
Association between early RfP and all the assessments of young adolescents across multiple domains related to behaviour and health. Behavioural and phenotypic assessments of young adolescents in the ABCD dataset were classified into 6 main categories, including physical health, cognition, mental (psychiatric) problems, substance use, culture and environment, and screen time (scales analysed in the study were listed in **Supplementary material 1)**. Each data point represents an *r* value derived from *LMM* association analysis between early RfP and a behaviour assessment subscale. If the *P* value passed Bonferroni correction, its *r* value was labelled as coloured points outside the horizontal dashed lines; otherwise, the *r* value was shown as grey-background points within the dashed lines. The scales that had the most significantly *r* values associated with early RfP were labelled and highlighted. For example, the highly positive *r* values of the cognition scales abcd_ps01 and abcd_tbss01 indicated that early RfP were highly positively associated with youths’ cognitive performance assessments, including subscales of the verbal learning (abcd_ps01) scale and dimensional cognitive summary (abcd_tbss01) scale. The 8 most significantly associated scales were as follows: Physical Health: **abcd_spacss01**, ABCD Sum Scores Parent-reported Sports and Activities Involvement on their children (including RfP-related scores); **(2)** Cognition: **absd_ps01**, ABCD Pearson Scores on verbal-learning and immediate-memory and **abcd_tbss01**, ABCD youth NIH TB cognition Summary Scores; **(3)** Mental Problems: **abcd_bpmt01**, ABCD Brief Problem Monitor Teacher-reported form, **abcd_cbcl01** ABCD parent-reported Child Behaviour Check List raw scores, **abcd_cbcls01** Child Behaviour Check List Summary scores, and **abcd_upps01** UPPS-P for Children Short Form (for impulsivity); and **(4)** Screen Time: **abcd_stq01** ABCD parent-reported Screen Time Survey.

Early RfP showed significant negative associations with young adolescent dimensional psychiatric assessments (Bonferroni-corrected *P* < 0.05), especially **1)** adaptive functioning and mental problems assessed by the CBCL (abcd_cbcl01, *r*: -0.034 ― -0.139), **2)** summary scale of psychiatric problems (abcd_cbcls01, *r*: -0.040 ― -0.105), **3)** brief problems monitor-reported by teachers (abcd_bpmt01, *r*: -0.047 ― -0.118) and **4)** UPPS-P impulsivity assessment (abcd_upps01, *r*: -0.031― -0.072). Interestingly, we found that early RfP was positively associated with youths’ sleep duration (abcd_sds01.sleepdisturb1_p, *r =* 0.068, *P* = 3.44×10^−13^) and negatively associated with their screen time (including time spent on TV or mobile-phones, etc.; abcd_stq01, *r*: -0.032 ― -0.092, Bonferroni-corrected *P* < 0.05).

The above results indicated that cognitive performance was better and mental problems were lower in young adolescents with early RfP. For the specific constituent subscales of cognitive and psychiatric assessments, we found that all 10 core standard subscales of NIH-TB cognition summary (abcd_tbss01) were positively associated with early RfP, while 11 out of the 20 core standard subscales of psychiatric problems summary (abcd_cbcls01) were negatively associated (**Figures 2A, B, Table S4**, Bonferroni-corrected). Among these subscales, the top-ranked positively associated cognitive subscales were: **1)** the crystallized composite of cognition (*r* **=** 0.331, *P* < 10^−20^) which was dependent upon past learning experiences(46), **2)** oral reading (*r* **=** 0.313, *P* < 10^−20^) and **3)** the total composite of cognition (*r* **=** 0.269, *P* < 10^−20^). Meanwhile, the top-ranked negatively associated psychiatric subscales were e: **1)** attention problems (*r* **=** -0.105, *P* < 10^−20^), **2)** ADHD symptoms CBCL-DSM-5 (*r* **=** -0.097, *P* < 10^−20^) and **3)** conduct problems (*r* **=** -0.083, *P* = 4.33×10^−17^). The total psychiatric scores (*r* **=** -0.069, *P* = 8.52×10^−12^) were also negatively associated with early RfP (**Figures 2C-H, Table S4**, Bonferroni-corrected). Other constituent subscales included positively associated picture vocabulary, fluid composite of cognition, etc., and negatively associated conduct, external and rule break psychiatric problems, etc. (**Figures 2A, B**, Bonferroni-corrected). Furthermore, similar findings were also observed in both the young adolescent male and female groups (**Table S5**, Bonferroni-corrected *P*<0.05).

**Figure. 2.**
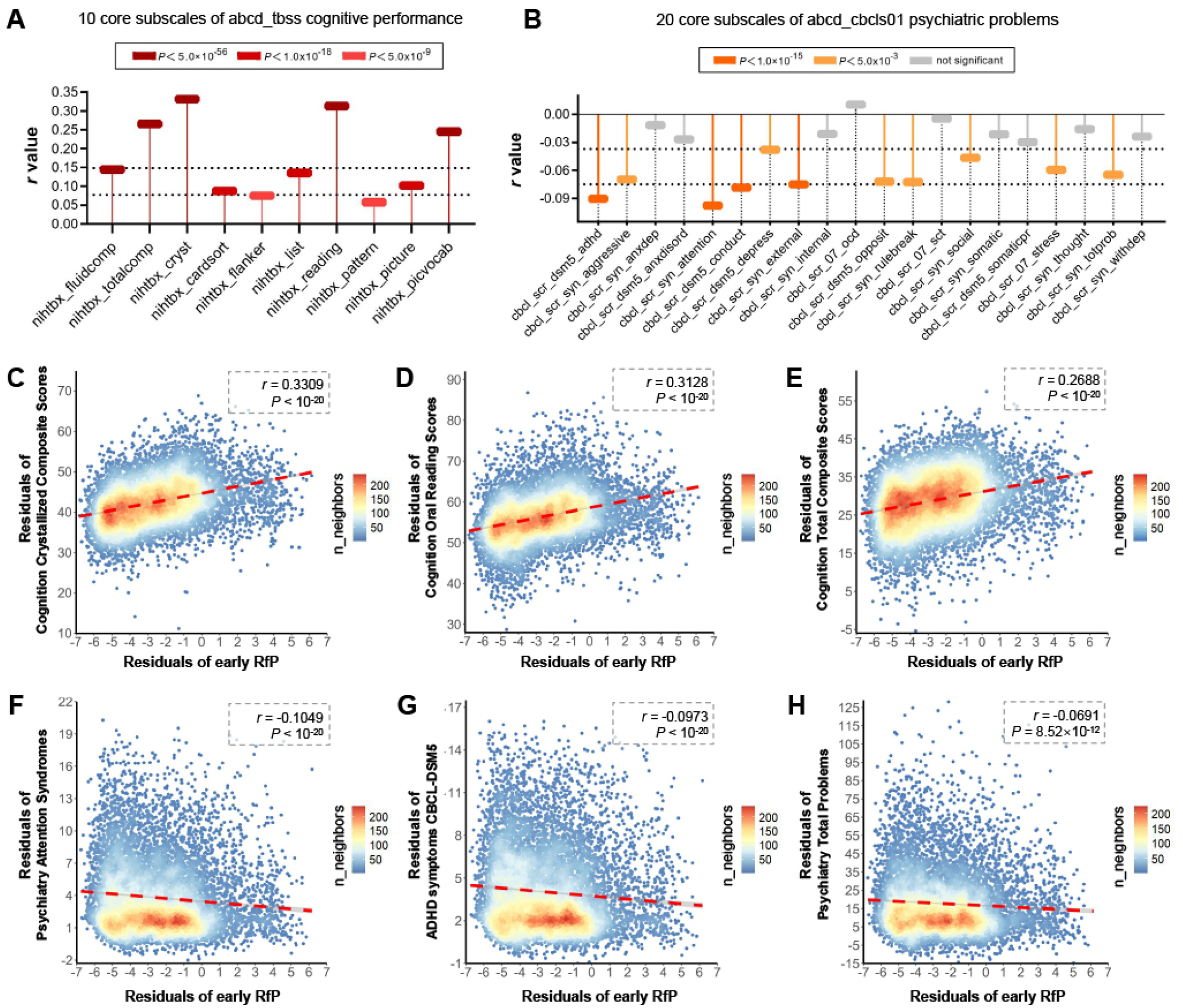
Early RfP, cognitive and psychiatric assessment scores in young adolescents. **(A)** The effect sizes of *LMM* associations between early RfP and all the core subscales of cognitive performance (abcd_tbss). **(B)** The effect sizes of *LMM* associations between early RfP and all the core subscales of psychiatric problems (abcd_cbcls01). **(C-E)** Density scatter plots showing the top 3 subscales of cognition that had the most significant positive associations with early RfP, including crystallized composite, oral reading and total cognition scores. **(F-H)** Density scatter plots showing the top subscales of psychiatric problems that had the most significant negative associations with early RfP, including attention problems, ADHD symptoms and total psychiatric problems. The analyses have regressed out all covariates. Each individual datapoint is coloured by the number of neighbouring points to represent the density of the overall data distribution. Bonferroni-corrected *P* <0.05. CBCL, child behaviour checklist; ADHD, attention deficit hyperactivity disorder.

### Mendelian randomization (MR) analysis of the effect between early RfP and later-life cognitive performance and ADHD

Using the IVW meta-analysis of MR based on SNP genetic instruments, we found a beneficial causal relationship between early RfP with better cognitive performance in adulthood (**Figure 3A, Table 2**, β = 0.0257, 95% CI: 0.0063 to 0.0451, *P* = 0.0094). The weighted median (WM) analysis was also significant (β = 0.0349, 95% CI: 0.0101 to 0.0597, *P* = 0.0059), and the MR–Egger results indicated a similar trend (β = 0.0543, 95% CI: -0.0324 to 0.1411, *P* = 0.2660). Steiger-MR directionality analysis confirmed the significant causal direction from early RfP to later-life cognition (**Table 2**, *P* < 10^−20^). A Forest plot showing effect sizes of single and combined SNPs indicated an all-combined positive effect pattern of early RfP on later-life cognitive performances (**Figure 3B**), and the causal estimates remained robust during the leave-one-out sensitivity test (**Figure 3C**), revealing no influence of any individual SNP on these results.

**Figure. 3.**
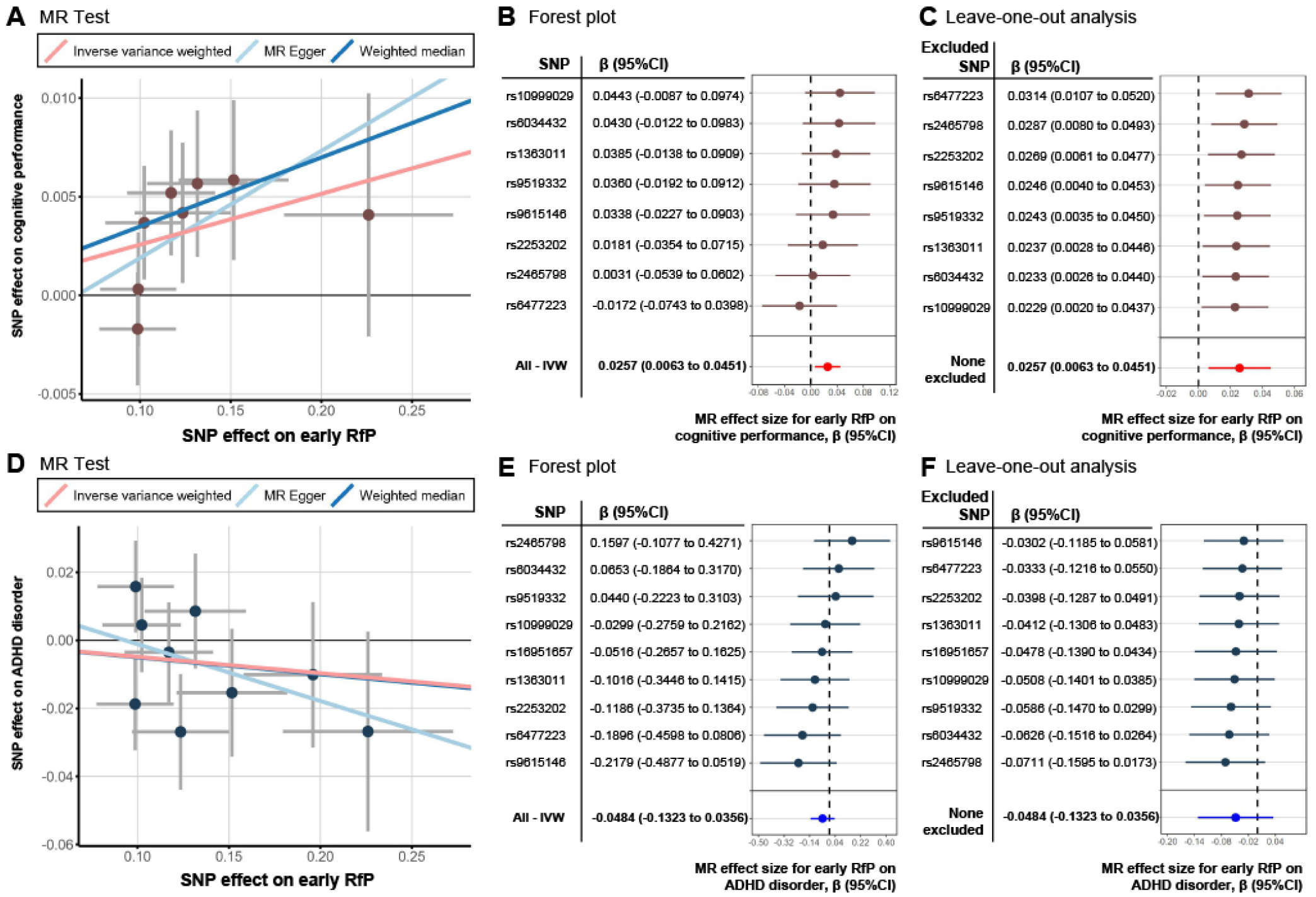
Effects of early RfP on later-life cognitive performances and attention syndromes analysed by 2-sample Mendelian randomization (MR). **(A)** Scatter plot showing the genetic instrument (SNP) effects on the exposure (early RfP) and the outcome (adult cognitive performance). The potential effects of the exposures on the outcomes using IVW, MR–Egger, and WM are shown by the regression lines, with the estimated effect represented by the slope. **(B)** Forest plot showing MR-analysed effect sizes of each single and all-combined SNP for the effect of early RfP on cognitive performance. **(C)** Leave-one-out sensitivity analysis that sequentially excluded each SNP from the estimation of the causal effect between of RfP on cognitive performance using the IVW method. **(D)** Scatter plot showing the SNP effects on the exposure (early RfP) and the outcome (adult ADHD). OR values had been converted to β statistics by log-transformation. **(E)** MR-analysed effect sizes of each single and all-combined SNP for the effect of early RfP on ADHD. **(F)** Leave-one-out sensitivity analysis for the estimation of the causal effect between early RfP on ADHD using the IVW method. IVW: inverse variance weighted, WM: weighted median.

**Table 2.** Results of Mendelian randomization (MR) analyses.

| The causal association of early RfP effect with later-life cognitive performances |  |  |  |  |  |  |
| --- | --- | --- | --- | --- | --- | --- |
| id.exposure | id.outcome | Method | No. of SNPs | $\beta$ | se | P Value |
| Early RfP | ebi-a-GCST006572<sup>a</sup> | IVW | 8 | 0.0257 | 0.0100 | **9.38×10<sup>-3</sup>** |
| Early RfP | ebi-a-GCST006572<sup>a</sup> | Weighted median | 8 | 0.0349 | 0.0127 | **5.87×10<sup>-3</sup>** |
| Early RfP | ebi-a-GCST006572<sup>a</sup> | MR Egger | 8 | 0.0543 | 0.0442 | 0.266 |
| The causal association of early RfP effect with later-life ADHD |  |  |  |  |  |  |
| id.exposure | id.outcome | Method | No. of SNPs | $\beta^c$ | se | P Value |
| Early RfP | ieu-a-1183<sup>b</sup> | IVW | 9 | -0.0484 | 0.0428 | 0.259 |
| Early RfP | ieu-a-1183<sup>b</sup> | Weighted median | 9 | -0.0498 | 0.0583 | 0.394 |
| Early RfP | ieu-a-1183<sup>b</sup> | MR Egger | 9 | -0.1670 | 0.1685 | 0.355 |
| Steiger MR directionality test |  |  |  |  |  |  |
| Early RfP → Later-life Cognition | id.exposure | id.outcome | snp_r2.exposure | snp_r2.outcome | correct_causal_direction | steiger_Pval |
|  | Early RfP | ebi-a-GCST006572<sup>a</sup> | 0.041 | 4.23×10<sup>-5</sup> | TRUE | 1.88×10<sup>-39</sup> |
| Early RfP → Later-life ADHD | id.exposure | id.outcome | snp_r2.exposure | snp_r2.outcome | correct_causal_direction | steiger_Pval |
|  | Early RfP | ieu-a-1183<sup>b</sup> | 0.047 | 1.42×10<sup>-4</sup> | TRUE | 9.74×10<sup>-41</sup> |

<sup>a</sup> Adult cognitive performances, <sup>b</sup> Children & adult ADHD disorder. <sup>c</sup> $\beta$ statistics were converted from OR by log-transformation in the ADHD case-control analysis. IVW, inverse variance-weighted analysis.

Regarding psychiatric problems, since there were no publicly available GWAS summaries associated with attention symptoms in normal adolescents/adults, we used SNPs associated with diagnosed ADHD in patients for the MR analysis. The results indicated a trend of protective negative causal relationship of early RfP with diagnosed children and adult ADHD disorder (β = -0.0484, 95% CI: -0.1323 to 0.0356, *P* = 0.2587) (**Figures 3D-F, Table 2)**. The Steiger-MR directionality test also confirmed the protective causal direction from early RfP to later-life diagnosed ADHD (**Table 2**, *P* < 10^−20^).

For sensitivity analysis of those MR evaluations, across all genetic instruments, no horizontal pleiotropy or heterogeneity was observed using the MR–Egger intercept test or modified Q statistics, and no SNP outliers were detected by the MR-PRESSO test.

### Brain structure is associated with early RfP, better cognitive performance and decreased psychiatric problems in young adolescents

Analysis with FreeSurfer and the standard Destrieux atlas revealed that the youths’ total brain volume (TBV) (**Figure 4A**) and intracranial volume (ICV) **(Table S6)** showed positive associations with early RfP. Interestingly, TBV and ICV were driven by brain growth during normal childhood development(5, 47, 48) and were decreased in psychopathology(49, 50). Youths with early RfP also showed increased specific brain cortical areas, including: **1)** bilateral: superior temporal and insula circular sulci, lateral superior temporal and middle frontal gyri; **2)** left-hemisphere (lh): angular, para-hippocampal, superior frontal and postcentral gyri, temporal pole, and middle frontal sulcus; and **3)** right-hemisphere (rh): anterior cingulate (ACC) region, middle temporal, middle occipital, supramarginal and orbital gyri (**Figure 4B, Table S6**, Bonferroni-corrected *P*<0.05). Early RfP was also positively associated with increased subcortical regions, including the ventral diencephalon (DC), thalamus, brain stem, and putamen regions (**Figure 4C, Table S6**, Bonferroni-corrected *P*<0.05).

**Figure. 4.**
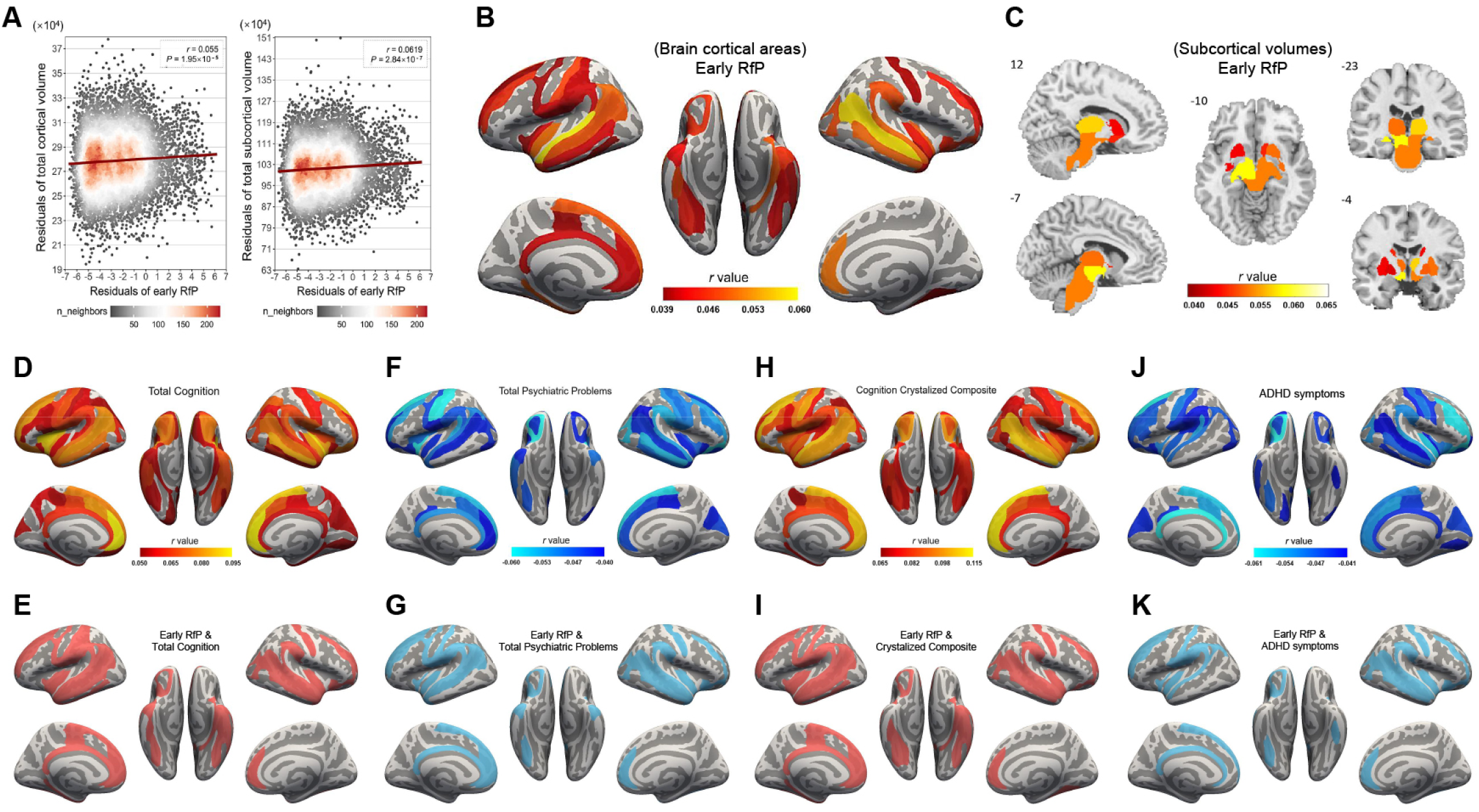
Young adolescent brain regions with their cortical areas and subcortical volumes significantly associated with early RfP, cognition, and psychiatric problems. **(A)** Density scatter plots showing that the total brain cortical volume (**left panel**, smri vol cdk total, mm^3^) and total subcortical volume (**right panel**, smri vol scs wholeb, mm^3^) were both positively associated with early RfP. **(B, C)** Brain map showing the specific cortical areas and subcortical volumes that were positively associated with early RfP. Regions with larger areas/volumes positively associated with early RfP are represented by the red colour. **(D, H)** Cortical areas that had significant positive associations with the total cognition score or crystallized composite cognition score. Regions where a large area was positively associated with a higher cognition score are represented by the red colour. **(F, J)** Cortical areas that had significant negative associations with total psychiatric problems and ADHD symptoms. Regions having a negative association between area and psychiatric assessment (i.e., a reduced cortical area was associated with increased ADHD symptoms) are represented by the blue colour. **(E, I)** All the increased cortical areas related to early RfP in **(B)** were overlapping regions that also positively correlated with the total cognition score **(E)** and crystallized composite score **(I). (G, K)** The overlapping brain regions with their areas positively associated with early RfP and negatively associated with the total psychiatric scores (**G**) as well as ADHD symptoms scores **(K)** are also shown. Bonferroni-corrected *P* <0.05.

The above early RfP-related structures were all important overlapping cortical and subcortical regions that were both significantly positively associated with the total and crystallized composite scores of cognition and negatively associated with total psychiatric problems and ADHD symptoms (**Figures 4D―K**, **Table S7**, Bonferroni-corrected *P*<0.05).

Validation analysis using the Desikan-Killiany atlas confirmed the above identified brain structures were related to early RfP (**Table S8**, Bonferroni-corrected *P*<0.05). Moreover, youths from both low- and high-income families had positive associations between early RfP and increased brain structure (**Table S9**, Bonferroni-corrected *P*<0.05).

Analysis of longitudinal structure data confirmed a significant positive association of baseline-year early RfP (ages 9–11) with brain structure recorded at the 2-year follow-up (ages 11–13) (**Table S10**, Bonferroni-corrected *P*<0.05).

Structural diffusion tensor (DTI) imaging analysis also revealed significant positive associations of early RfP with white-matter fibre tract volumes of the whole brain and specific brain regions, including the corticospinal/pyramidal, temporal and superior longitudinal fasiculus, fornix, and inferior-fronto-occipital and anterior thalamic radiations (**Table S11**, Bonferroni-corrected *P*<0.05)

### The longitudinal association and mediation analyses

Using Structural equation modelling, we analysed the changes of assessment scores between the baseline year and 2-years later, with confounders regressed. Consistent with the MR findings, the longitudinal analysis confirmed that early RfP levels recorded at baseline year were significantly associated with improved cognitive performance (β = 0.080, *P* < 10^−4^) and reduced ADHD symptoms (β = - 0.029, *P* = 0.009) at the 2-year follow-up **(Figures 5A, B)**.

**Figure. 5.**
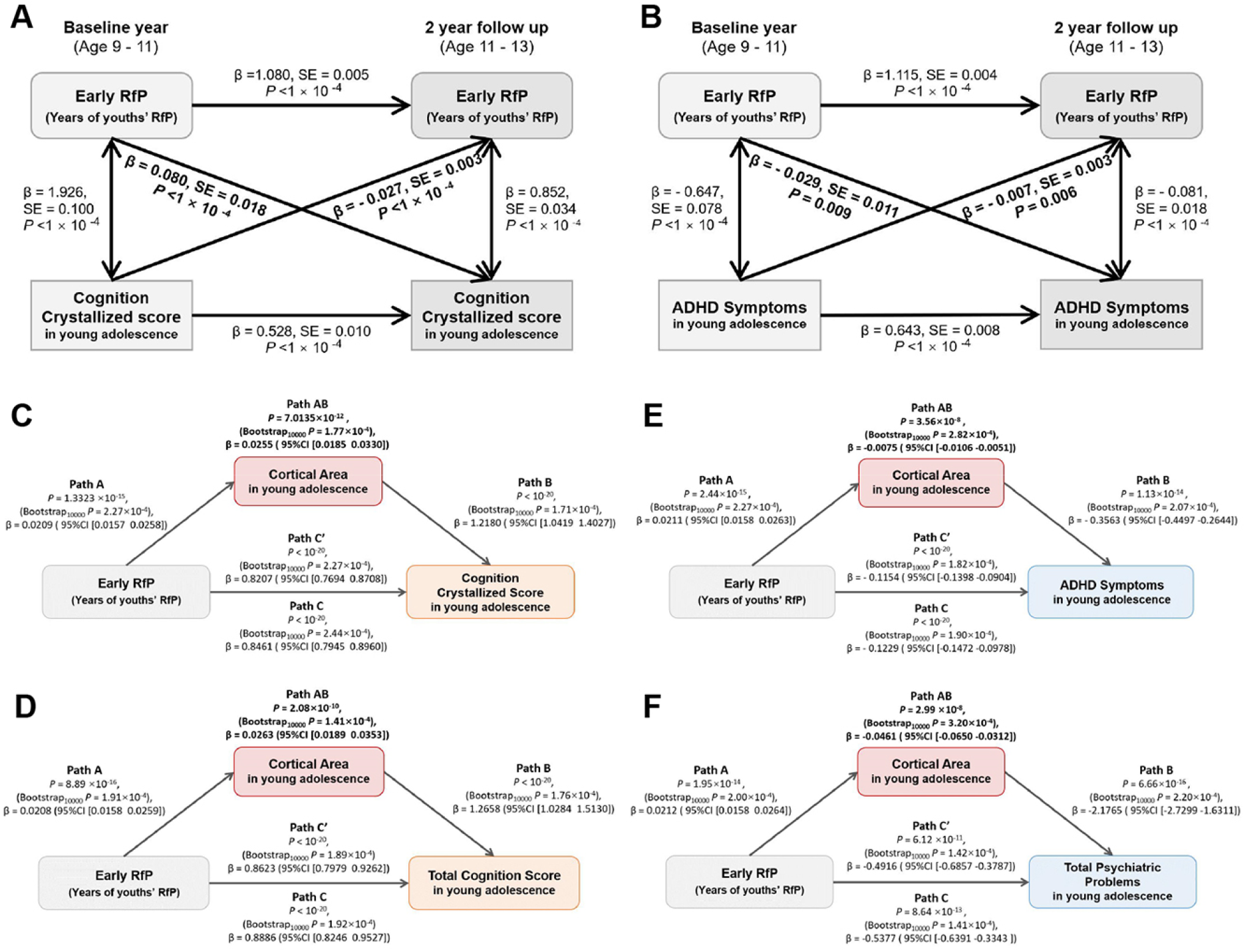
The longitudinal association and mediation analysis between early RfP and cognitive performance/psychiatric problems. **(A, B)** The structural equation modelling analysis indicated a longitudinal relationship of the early RfP with cognitive score as well as attention problems (the CLPM model was used). Higher scores of early RfP were associated with higher cognitive assessment scores (β = 0.080, SE = 0.018, *P* < 1.0 × 10^−4^) and lower attention symptoms (β = -0.029, SE = 0.011, *P* = 0.009) measured 2 years later. **(C-F)** Path A indicates the direct effect of the predictor factor (early RfP) on the mediator (brain cortical area in young adolescence); Path B indicates the direct effect of the mediator on the behaviour assessment outcome (young adolescent cognitive performance/psychiatric problem); Path C indicates the total relationship of the predictor factor on the behaviour assessment outcome. path C’ indicates the direct effect, controlling for the mediator. Path AB is the product of path A and path B (βA*\**βB), indicating the mediation effect of the predictor factor on the behaviour assessment through the cortical area. The path AB effects implemented by cortical structures from early RfP on cognitive performance/psychiatric problems were all significant (*P* and bootstrap *P* <0.001). The β values represent regression coefficients of the effect of the independent variables on the dependent variables. CLPM: two-wave cross-lagged panel model.

The significant mediation pathway that linked the association between early RfP and youths’ cognitive performance was the path AB of cortical area (for the crystallized composite score: β = 0.0255, 3% of the total effect size, *P* = 7.01×10^−12^, 95% CI: 0.0185 to 0.0330, **Figure 5C**; for total cognition score: β = 0.0263, 3% of the total effect size, *P* = 2.08×10^−10^, 95% CI: 0.0189 to 0.0353, **Figure 5D**). A similar finding was also observed for psychiatric problems, which indicated a significant mediation effect of the cortical area (for ADHD symptoms: β = - 0.0075, 6% of the total effect size, *P* = 3.56×10^−8^, 95% CI: - 0.0106 to - 0.0051, **Figure 5E**; for total psychiatric scores: β = - 0.0461, 8.6% of the total effect size, *P* = 2.99×10^−8^, 95% CI: - 0.065 to -0.0312, **Figure 5F)**. Mediation analysis of subcortical brain volumes also showed significant mediation effects **(Figure S2**, *P* < 10^−4^). Our hypotheses were confirmed by the analysis that the brain structure changes of specific cortical areas and subcortical volumes in **Figures 4E, G, I**, and **K** and **Table S7** significantly mediated the relationships of early RfP with youths’ cognitive performance, as well as with psychiatric problems (**Figure 5C-F, Figure S2**).

### Beneficial level of young adolescent RfP durations were significantly associated with cognition, mental health and brain structure

Scatter-plot and nonlinear fit models indicated that the optimal weekly RfP duration of youths for cognitive performances was approximately 12 hours (h)/week, representing the beneficial RfP habits **(Figures 6A, B)**. Under the condition of within 12 h/week RfP, the increasing rates of cognitive scores accompanied by the growth in RfP durations were most notable, indicated by significant correlations (crystallized composite: *r* = 0.323, *P* < 10^−20^; total cognition: *r* = 0.253, *P* < 10^−20^). However, in conditions of more than 12 h/week RfP, no additional benefits but slow decreases in crystallized composite (*r* = -0.137, *P* =0.025) and total cognition (*r* = -0.125, *P* =0.083) scores were observed accompanied with increases in RfP durations. It’s possible that overextended time of RfP might be detrimental since it possibly reduces time spent in other daily activities known to have benefits on cognition, including sports, schooling and social activities. We further observed moderate linear negative correlations of RfP durations with psychiatric problems, including attention (*r* = -0.045, *P* = 3.53×10^−4^) and CBCL-DSM-5 conduct problems (*r* = -0.0375, *P* = 0.007) **(Figures S3 A, B** Bonferroni-corrected).

**Figure. 6.**
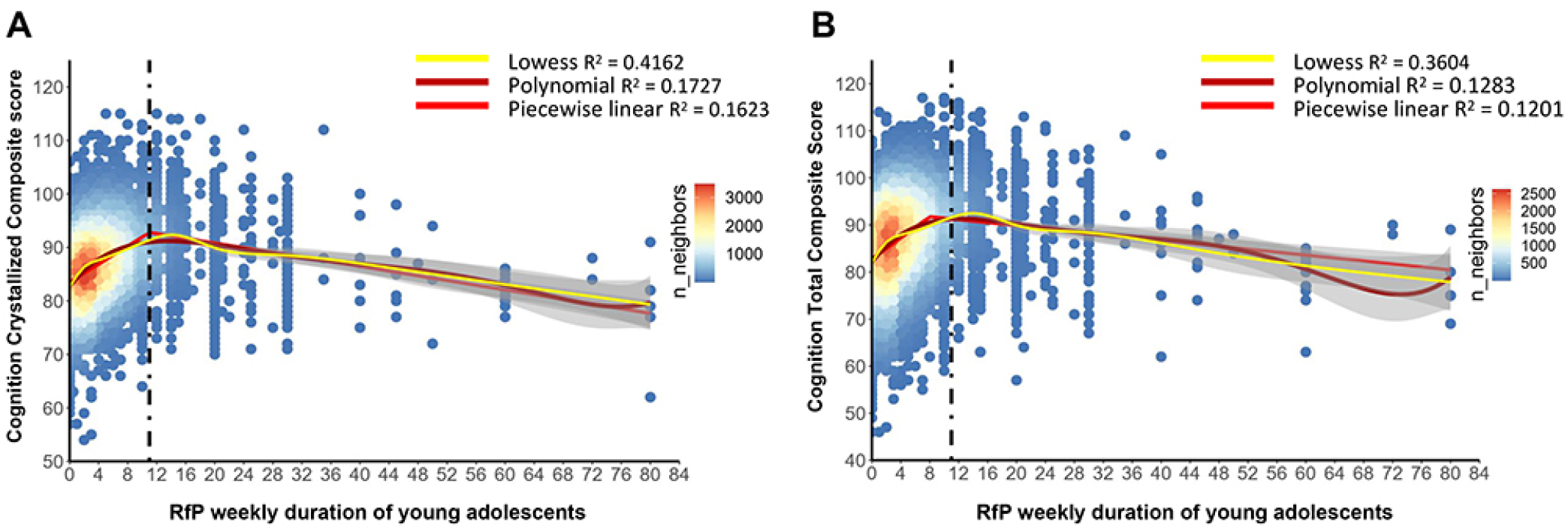
Nonlinear associations between RfP durations and cognitive performance of young adolescents. **(A, B)** Scatter plots showing the association between RfP habits (represented by appropriate RfP hours(h)/week) of young adolescents and their cognitive scores. Nonlinear fit results indicated that the optimal RfP duration for cognitive performance, including the cognition crystallized composite and total cognition score, was approximately 12 h/week, as represented by the black dotted line. For the condition of less than or equivalent to 12 h of RfP per week, the cognition assessment scores improved with increasing RfP time. Bonferroni-corrected *P* <0.05. For the condition of more than 12 h RfP per week, cognition scores decreased slowly with increasing RfP time.

Youths who maintained beneficial RfP durations also had increased cortical volumes of the brain regions defined above, such as the temporal, insular, superior frontal, and subcortical ventral DC, putamen, thalamus and brain stem (**Table S12**, Bonferroni-corrected *P*<0.05), while in conditions of more than RfP for 12 h/week, no significant changes in brain structure was observed in association with increased RfP durations.

Longitudinal analysis suggested that beneficial RfP durations (within 12 h/week) at baseline were significantly associated with improved cognitive performance at the 2-year follow-up (β = 0.205, *P* < 10^−4^), and the reverse was also significant (β = 0.082, *P* < 10^−4^); however, baseline-year ADHD symptoms were significantly associated with decreased beneficial RfP durations 2-year later (β = - 0.025, *P* = 0.004), and the reverse was not significant in young adolescents **(Figures S4 A, B)**. Mediation analysis demonstrated that the beneficial RfP duration significantly mediate the relationship between cortical structure and cognitive performance (β = 0.2403, 15.3% of the total effect size, *P* = 5.15×10^−11^, 95% CI: 0.1433 to 0.2659); additionally, lower ADHD symptoms significantly mediate the effect of increased brain structure on increased beneficial RfP durations (β = 0.0248, 9.2% of the total effect size, *P* = 1.47×10^−7^, 95% CI: 0.0161 to 0.0356) in young adolescence **(Figures S4 C, D)**.

## Discussion

There has been a widespread consensus that exposure to publications brings us more wisdom and helps promote more beneficial outcomes in life. Caregivers were encouraged to assist young children’s reading(51, 52), and children were also encouraged to read by schools, governments and social media(21). Here, we performed for the first time a large-scale analysis on the attainments of early childhood reading for pleasure (early RfP) during the important initial development stages of youths and found unique evidence of it in laying beneficial cognitive, mental, and brain structural foundations for later-learning and well-being.

We found positive associations between early RfP and youths’ higher cognitive test scores, which showed improvements in dimensional cognitive performance covering major neurocognitive domains, regressing out the family SES(53) statuses. The results demonstrated that early RfP had similar beneficial associations with cognition as educational attainments (EA), since EA were positively associated with cognitive function in later-life(19), which might partly be due to cognitive enrichments(54-56). Our findings extend the importance of formal education to an enjoyable early educational activity during the critical formative stage. Furthermore, we found negative associations between early RfP and youths’ attention problems, consistently, a previous observational study using the UK Millennium Cohort (MCS) had suggested a link between RfP frequencies in children at age 7 and lower inattention at age 11(57). Additionally, our findings extended the associations with a broad range of reduced psychiatric problems, including aggressive, conduct, external, rule-break, stress problems.

We further performed MR analyses using genetic instruments obtained from independent large-scale GWAS summaries and found a potential causal relationship of early RfP with better cognitive performance and a trend of protective relationship with lower ADHD in later life. Our findings were consistent across different established MR methods. Although the molecular and neurobiological mechanisms through which the SNPs affect early RfP were unclear, the SNPs with known gene consequences were found to be closely related to neuronal functions. For example, rs2253202 is related to the KCNMA1 gene, which is fundamental to neuronal excitability control. Mutation of the gene causes developmental delay and seizures(58). Additionally, rs10999029 is related to the synaptic extracellular-matrix protein COL13A1, which is involved in neuromuscular synapses(59). Similar to early RfP, EA had positive causal effects on cognition(19, 60). EA also impacts alcohol consumption problems, such as reducing the frequency of binge drinking and reducing AD risk(61).

We found that early RfP was significantly associated with youths’ cortical structures, especially the temporal, frontal, angular, parahippocampal, ACC, middle-occipital, supramarginal and orbital regions. Some of these regions were found to play important roles in reading-related processes: a study of neurodevelopmental mechanisms of reading(62) showed that the brain temporoparietal cortices, containing the left superior temporal gyrus, matured early in learning and remained involved in reading throughout adulthood. Specifically, the left superior temporal is engaged in early reading acquisition and is related to children’s phonological awareness skills(62, 63). The superior temporal functioned critically in cross-modal integration, such as processes associated with the mapping of print to sound(62). sMRI revealed that the literate group had greater grey matter density in the bilateral middle temporal and dorsal occipito-parietal, left superior temporal and supramarginal regions(64) than the illiterate groups. Consistently, brain regions, including the angular, temporal and supramarginal gyri, are activated during reading, and their grey matter density increases were induced by literacy acquisition(65).

We further identified that brain regions positively related to early RfP were overlapping regions both positively associated with cognitive performance and negatively associated with psychiatric problems, among which, the temporal, frontal, cingulate and orbital regions played key roles in neuronal processes related to attention, emotional processing, reward/punishment, response inhibition, neurodevelopmental and psychiatric disorders(66-70). Specifically, the temporal(71), prefrontal(70), and ACC(72) cortices play important roles in cognitive functions, and the orbitofrontal cortex is involved in depressive and related mental problems(68, 73). These RfP-related regions are also known to be impaired in ADHD(74, 75). A recent large-scale study in paediatric populations confirmed that the frontal, temporal, and cingulate regions of children with ADHD showed decreased cortical areas(75). Analysis across different cohorts confirmed that ADHD patients had decreased ICV and subcortical regions, including the accumbens, caudate, and putamen(49).

We also found positive associations of youth total brain TBV and ICV with early RfP. Fundamentally, children’s TBV and ICV develop in parallel until young adolescence, after which TBV gradually declines while ICV becomes stabilized(5, 47, 48). Reduced TBV/ICV was found in patients with ADHD(49) and schizophrenia(50), which might be caused by disruption of early brain development or underdevelopment (i.e., hypoplasia)(76).

Future studies should include an investigation of the long-term persistence of these associations. Longitudinal analyses from young adolescence to adulthood should be conducted on RfP measurements in subsequent updates of the ABCD project.

RfP in all children may support the best possible chance for good cognitive and mental development. A relatively simple early educational initiative, reading enjoyable children’s book/publications for pleasure, could have beneficial associations with cognition, performances in school, the brain, and mental health in young adolescence and possibly even beyond. Therefore, the findings have valuable implications for caregivers, teachers and policy makers globally(77).

## Data Availability

This study was conducted under the ABCD NDA data permission number of 9143 (DAR ID).

## Acknowledgements

We thank all the subjects and their families for participating in the ABCD project and for their time and dedication. We acknowledge that the behavioral and neuroimaging data used in this study were obtained from the ABCD project (https://abcdstudy.org). The ABCD Project is supported by the NIH and additional federal partners. A list of all supporters can be found at https://abcdstudy.org/federal-partners.html.

This research was supported by the following grants:

Dr Sahakian receives funding from a Wellcome Trust Collaborative Award (200181/Z/15/Z), and the research is conducted within the NIHR Cambridge Biomedical Research Centre (BRC) (Mental Health Theme and Neurodegeneration Theme) and the NIHR MedTech and in vitro diagnostic Co-operative (MIC).

Dr Feng was supported by National Key R&D Program of China (No. 2019YFA0709502), National Key R&D Program of China (No. 2018YFC1312904), Shanghai Municipal Science and Technology Major Project (No. 2018SHZDZX01), ZJ Lab, Shanghai Center for Brain Science and Brain-Inspired Technology, and the 111 Project (No. B18015).

Dr Cheng was supported by grants from the National Natural Sciences Foundation of China (No. 82071997), the Shanghai Rising-Star Program (No. 21QA1408700) and Natural Science Foundation of Shanghai (No. 18ZR1404400).

Dr Sun was supported by grants from the National Natural Science Foundation of China (No. 31700892).

The funders had no role in study design, data collection and analysis, decision to publish or preparation of the manuscript.

